# Bleeding Risk with Combination Antithrombotic Therapy: A Propensity-Score Matched Analysis

**DOI:** 10.1101/2025.01.07.25320165

**Authors:** Salia Farrokh, Krishna Nalleballe, Sanjeeva Onteddu, Jose I. Suarez, Julian Bösel, Vishank A. Shah

**Affiliations:** Division of Neurosciences Critical Care, Department of Anesthesiology and Critical Care Medicine, Neurology, and Neurosurgery, Johns Hopkins University School of Medicine, Baltimore, MD; Department of Neurology, University of Arkansas for Medical Sciences, Little Rock, AR; Department of Neurology, Friedrich-Ebert-Krankenhaus (FEK), Neumünster, Germany; Department of Neurology, University Hospital Heidelberg, Heidelberg, Germany

**Keywords:** Anticoagulation, Antiplatelet therapy, Acute ischemic stroke, Acute myocardial infarction, Atrial fibrillation, Intracerebral hemorrhage risk, Acute gastrointestinal bleeding risk

## Abstract

**Background:** Limited data exist on the safety of combining antiplatelets and anticoagulants (AC) for secondary stroke prevention in acute ischemic stroke (AIS). We sought to examine the hemorrhage risk with combining single (SAPT) or dual antiplatelet therapy (DAPT) with AC in AIS patients with concomitant atrial fibrillation (AF) and acute myocardial infarction (MI).

**Methods:** This retrospective cross-sectional cohort study used TriNetX, a federated health analytics database that collects real-time electronic health records data from 76 participating healthcare organizations in the United States. Through queries performed on March 31, 2023, adult patients with simultaneous diagnoses of AIS, AF and acute MI were identified using ICD-10 codes over the past 20 years. Propensity score-matched analysis, matching for demographics and vascular co-morbidities, compared the odds for acute spontaneous ICH and GI bleeding at 3-months, 12-months and throughout follow-up within TriNetX, between three matched sub-cohorts: AC alone, AC+SAPT, and AC+DAPT.

**Results:** Among 144,434 AIS patients with AF and MI (mean age, 71.9 years; 43.3% female), 8772 (6.1%) patients received AC alone, 88,430 (61.2%) received AC+SAPT, and 47,232 (32.7%) received AC+DAPT. After propensity-score matching, 8,706 patients were included in each sub-cohort. Compared to AC alone, AC+SAPT and AC+DAPT showed no significant increase in ICH risk at 3 and 12 months but increased long-term risk throughout follow-up (odds ratio [95% confidence intervals] for AC+SAPT, 1.26 [1.11-1.44], p<0.001; for AC+DAPT, 1.34 [1.18-1.53], *p*<0.001). Gastrointestinal (GI) bleeding risk was elevated at all time-points with combination therapies.

**Conclusion:** Within the limitations of a retrospective cross-sectional study using administrative data, this large propensity-score matched analysis demonstrates that combining antiplatelets with anticoagulants after AIS may be associated an increased risk of ICH only in the long-term, beyond 12-months, but with an increased risk of acute GI bleeding in the short and long-term. Larger prospective studies are warranted to confirm our findings.

## Introduction

Non-valvular atrial fibrillation (AF) is a major risk factor for acute ischemic strokes (AIS). When stroke occurs in patients with AF, it is often more severe and is associated with a higher risk of disability and mortality than those without AF (HJ 1996). According to clinical guidelines, anticoagulation (AC) reduces the risk of cardioembolic AIS in patients with AF and therefore those at high risk indicated by CHA_2_DS_2_-VASc score of 2 or more should receive AC indefinitely for ischemic stroke prevention (GYH 2010). In fact, the superiority of AC therapy over aspirin for stroke prevention in patients with nonvalvular AF is well-established (Lip & GYH 2006).

Patients with AIS and atrial fibrillation (AF) often have a comorbid history of coronary artery disease (CAD) in up to 25% to 35% of cases (Nabauer, 2009; Steinberg, 2013) and may present with concomitant acute myocardial infarction (MI) in up to 1% to 2% of cases (Alqahtani, 2017). Coronary artery disease, particularly in the setting of acute MI, warrants single (SAPT) or dual antiplatelet therapy (DAPT) with low-dose aspirin and a P2Y12 inhibitor agent such as clopidogrel, prasugrel, or ticagrelor (Amsterdam 2014). Moreover, according to the 2021 AHA guidelines for CAD, patients require DAPT for 1-6 months following percutaneous coronary intervention (PCI) before de-escalation to single antiplatelet therapy with a P2Y12 inhibitor (O’Gara 2021). This highlights the clinical dilemma of which therapeutic strategy should be pursued for patients who require AC therapy for cardioembolic stroke prevention but also require antiplatelet therapy for immediate cardiovascular protection.

Often SAPT is combined with anticoagulation in patients with AF and CAD, particularly in the setting of acute MI. For example, the American College of Cardiology (ACC) guidelines recommend combining a direct oral anticoagulant (DOAC) and a P2Y12 inhibitor for the first 6-12 months after PCI followed by AC monotherapy thereafter (Kumbhani 2021). Additionally, the ACC guidelines provide four clinical scenarios where the use of triple antithrombotic therapy may be warranted: 1) patients receiving AC for AF who undergo PCI and require DAPT 2) patients who are already receiving DAPT for atherosclerotic cardiovascular disease (ASCVD) with newly diagnosed AF who require AC for stroke prevention 3) patients who are receiving AC for venous thromboembolic disease (VTE) and need PCI and therefore require addition of DAPT 4) patients on DAPT for ASCVD and newly diagnosed VTE who require AC (Kumbhani 2021). According to these guidelines, the duration of triple therapy should be limited to 30 days and likely needs to be individualized based on individual patient risk factors (Kumbhani, 2021). Prior reports show that triple therapy significantly increases the risk of bleeding; the reported bleeding risk when adding SAPT to AC therapy is about 20%-60% compared to adding DAPT to oral AC, which is estimated to be 2 to 3-fold (Dentali 2007, Hansen 2010, Lopes 2019, Dewilde 2013, Gibson 2016, Cannon 2017, Vranckx 2019). More recently, the Effective Anticoagulation with Factor Xa Next Generation in Atrial Fibrillation–Thrombolysis in Myocardial Infarction (ENGAGE AF-TIMI) 48 trial showed that concomitant use of SAPT and AC resulted in similar incidence of ischemic events but was also associated with higher risk of life-threatening systemic and intracranial bleeding (Xu 2016). Despite this, patients are often prescribed long-term combination therapy, with 12-26% receiving lifelong combination of an antiplatelet agent with AC (Fox 2020, Lamberts 2014).

This clinical conundrum is further embellished in AIS survivors, where the risk of intracerebral hemorrhage in the acute setting and long-term may be 15 times higher than patients without an AIS (Ögren, 2015). However, the majority of the highlighted studies evaluating combining antiplatelets and AC in AF, particularly triple antithrombotic therapy, do not include patients with AIS. Therefore, limited data exist regarding the frequency of combining antiplatelets with anticoagulation in current clinical practice and the safety of such combination therapies in AIS survivors.

The objective of this study was to describe the frequency of combination antiplatelet and anticoagulant prescriptions in clinical practice and define the short-term and long-term risk of intracerebral hemorrhage (ICH) and acute gastrointestinal (GI) bleeding when combining antiplatelets and AC therapy in AIS patients with AF and concomitant acute MI. We hypothesized that DAPT or SAPT in combination with AC would be associated with a higher risk of ICH and GI bleeding in both the short and long-term after AIS.

## Methods

### Study Design, Data Source and Availability

We conducted a retrospective cross-sectional cohort study on de-identified patient data included in the TriNetX database. TriNetX is a global federated health research network recording electronic health records (EHR) data in real-time from a large network of participating academic and community healthcare organizations located mainly in the United States (US) and US territories (TriNetX, n.d). Data included in the TriNetX platform include patient demographics, diagnoses classified using the *International Classification of Diseases, Tenth Revision, Clinical Modification* (ICD10-CM) codes, individual medication data and medication anatomic Therapeutic Chemical codes, among other data (TriNetX, n.d).While TriNetX collects data from the EHR in real-time, leading to no lag between hospital admission and data entry, the de-identification of the data limits access to details regarding hospital site name, size, hospital characteristics or precise location. However, due to the availability of large volumes of patient-level data, linked to billing codes, TriNetX provides valuable data for comparative effectiveness analyses and has been previously used in several other clinical studies (Bucci, 2024, Shah 2020, Alobaida, 2022). We obtained Institutional Review Board approval from the University of Arkansas for Medical Sciences prior to conduct of this study, with exemption for consent given that study involved de-identified data from a healthcare analytics platform that is compliant with the Health Insurance Portability and Accountability Act (HIPAA) and the US federal law which protects the privacy and security of healthcare data. We adhered to the Strengthening the Reporting of Observational Studies in Epidemiology (STROBE) guidelines for reporting of this study (Elm, 2020). Datasets can be accessed through queries performed on the TriNetX platform (TriNetX, n.d).

### Study Cohorts

For this analysis, cross-sectional cohorts were queried by one physician (K.N.) on March 31, 2023, using browser and real-time features on a TriNetX network called “Research”, which included 76 participating healthcare organizations across US and US territories, of which 52 centers responded with patients that met inclusion criteria for this study. Inclusion criteria were (i) adult patients (≥18 years of age); (ii) primary discharge diagnosis of acute ischemic stroke (AIS) or cerebral infarction (ICD10-CM: I63); (iii) underlying diagnostic code for atrial fibrillation or flutter (AF) (ICD10-CM: I48); and (iv) a simultaneous new diagnostic code for acute myocardial infarction (MI) (ICD10-CM: I21). We did not include a separate code for percutaneous coronary interventions (PCI) to maximize patient retention in the cohorts given that acute MI patients often receive antiplatelet therapy even in the absence of PCI (Levine, 2016). To further maximize patient inclusion, we did not limit our search to a specific time window, however none of the patients included had an index event more than 20 years prior to the query date (i.e., March 2003), an approach that has been used previously (Bucci, 2024).

Patients were categorized into 3 sub-cohorts based on medication exposure: (i) Group 1: AC alone (medication code, NLM:VA:BL110; ICD10-CM code, Z79.01); (ii) Group 2: AC plus SAPT (above code plus medication [code] for Aspirin [NLM:RXNORM:1191] or Clopidogrel [NLM:RXNORM:32968] or Ticagrelor [NLM:RXNORM:1116632]); (iii) Group 3: AC and DAPT (medication codes for AC plus any two antiplatelet agent medication codes). Comparisons were made between patients with AC alone versus AC+ SAPT and AC alone versus AC+DAPT.

### Outcomes

The primary outcome for the study was risk of non-traumatic spontaneous ICH (ICD10-CM code: I61) and secondary outcome was the risk of gastrointestinal bleeding (GIB) (ICD10-CM: K92.2). Outcomes were measured at three time points: (i) within 90 days; (ii) within 12 months; and (iii) at any timepoint after the index event until the query date.

### Statistical Analysis

All analyses were conducted using the TriNetX research analytics platform that uses R package 2.0. Baseline characteristics, including demographics and medical comorbidities were compared using the independent sample’s *t*-test for continuous variables and χ^2^ test for categorical variables. To generate balanced cohorts, the TriNetX analytics platform was used to generate individual propensity scores using logistic regression, after which 1:1 propensity-score matching was performed utilizing ‘greedy nearest neighbor’ algorithms with a pre-specified caliper width of 0.1 pooled standard deviations (SD), methodology that has been used previously in multiple studies (Bucci 2024, Alobaida 2023, Decker 2022). Propensity score density graphs were examined and a standardized mean difference of <0.1 between cohorts was considered as well-balanced. Cohorts were matched using the following patient characteristics: age, race, sex, ethnicity, hypertension, type 2 diabetes, heart failure, coronary artery disease, chronic obstructive pulmonary disease, asthma, smoking, chronic kidney disease, obesity, and hyperlipidemia, classified using ICD10-CM codes (Supplement for details on ICD10-CM codes used). These characteristics were chosen given their clinical relevance to cardiovascular disease burden, short- and long-term mortality and morbidity risk after AIS. logistic regression was used in propensity matched cohorts to assess odds for ICH and acute GIB at the pre-specified timepoints after the index event.

## Results

### Cohort Characteristics

Within TriNetX, 144,434 patients were identified with an index event of AIS and concomitant acute MI as well as AF using ICD10-CM diagnostic codes. The mean (SD) age of the entire cohort was 71.9 (12.3) years, 62,521 (43.3%) were female; 110,537 (76.5%) were White, 17,921 (12.4%) Black, 1700 (1.2%) Asian and 5906 (4.1%) of Hispanic Ethnicity. Among the entire cohort, 8772 (6.1%) patients received AC alone, 88,430 (61.2%) received AC + SAPT, and 47,232 (32.7%) patients received AC + DAPT ***(Figure 1)*.**

**Figure 1:**
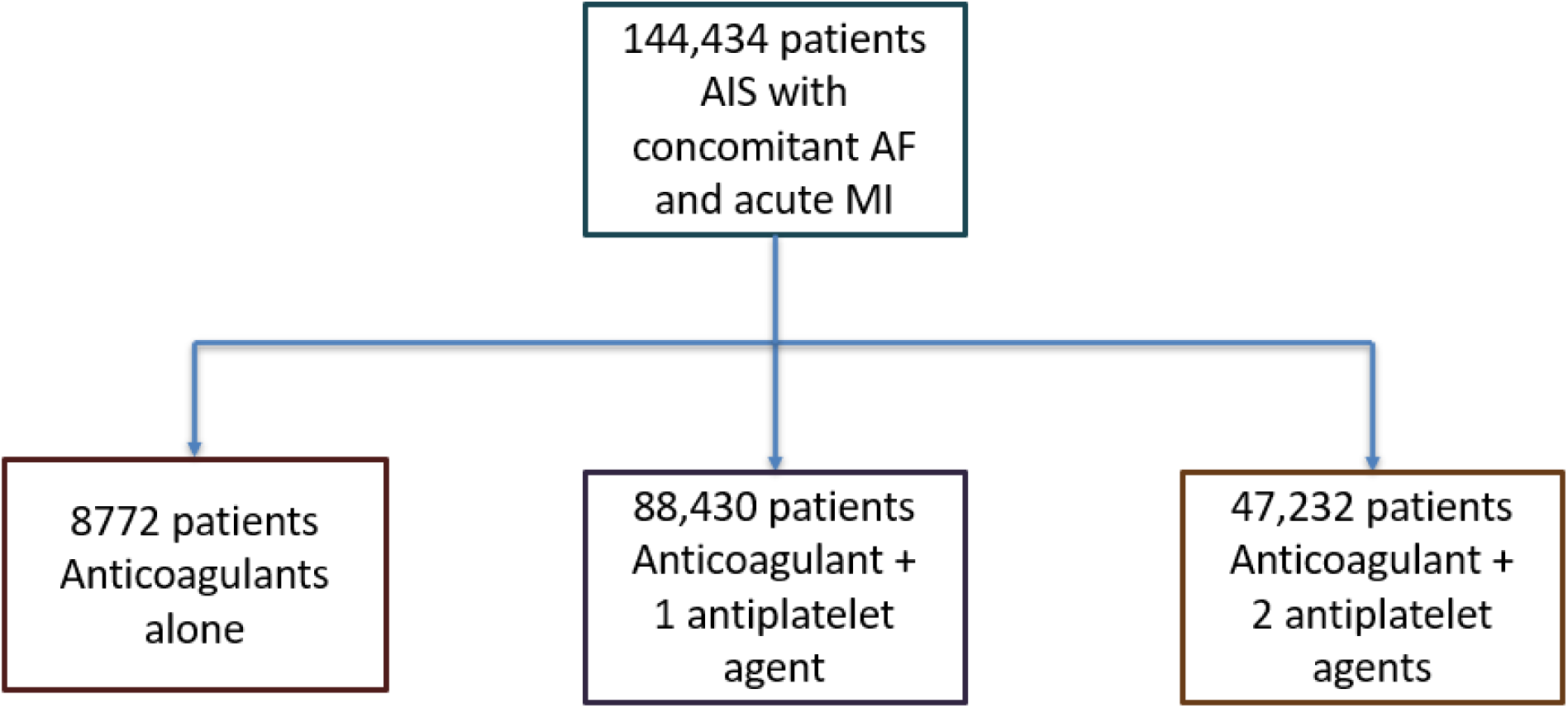
Patient Selection Flow Diagram. This figure shows how patients were grouped in different study sub-cohorts: anticoagulants alone, anticoagulants and 1 antiplatelet agent, anticoagulant and 2 anticoagulant agents. **Abbreviations:** AIS, Acute Ischemic Stroke; AF, Atrial Fibrillation; MI, Myocardial Infarction

Comparison of baseline characteristics between the groups are listed in *Table 1*. Compared to group 1, more patients in group 2 and 3 had significantly higher prevalence of medical co-morbidities including type 2 diabetes, coronary artery disease, and heart failure, among others *(Table 1)*.

**Table 1:** Baseline Characteristics.

| Variable | AC alone<br>(N=8772) | AC + SAPT<br>(N=88,430) | AC + DAPT<br>(N=47,232) | P-value (AC<br>alone vs. AC<br>+ SAPT) | P value (AC<br>alone vs.<br>AC +<br>DAPT) |
| --- | --- | --- | --- | --- | --- |
| Age, mean (SD) y | 72.1 (14.0) | 71.8 (12.4) | 72.0 (11.6) | 0.03 | 0.18 |
| Race/Ethnicity, n<br>(%) | 5594 (64.3) | 68,170 (77.4) | 36,773 (78.1) | <0.001 | <0.001 |
| White | 955 (11.0) | 11,273 (12.8) | 5693 (12.1) | <0.001 | 0.003 |
| Black | 102 (1.2) | 1027 (1.2) | 571 (1.2) | 0.96 | 0.75 |
| Asian | 311 (3.6) | 3624 (4.1) | 1971 (4.2) | 0.02 | 0.008 |
| Hispanic |  |  |  |  |  |
| Female Sex, n (%) | 4269 (49) | 38,700 (43.9) | 19,552 (41.5) | <0.001 | <0.001 |
| Comorbidities, n<br>(%) | 6905 (79.3) | 80,065 (90.9) | 43,752 (92.9) | <0.001 | <0.001 |
| Hypertension | 3785 (43.5) | 52,719 (59.8) | 30,066 (63.8) | <0.001 | <0.001 |
| Type 2 Diabetes | 1477 (17.0) | 18,662 (21.2) | 11,626 (24.7) | <0.001 | <0.001 |
| Hyperlipidemia | 2410 (27.7) | 34,857 (39.6) | 19,554 (41.5) | <0.001 | <0.001 |
| Obesity | 4369 (50.2) | 64,393 (73.1) | 38,748 (82.3) | <0.001 | <0.001 |
| CAD | 4247 (48.8) | 53,706 (61.0) | 29,888 (63.5) | <0.001 | <0.001 |
| Heart Failure | 3479 (40.0) | 43,221 (49.1) | 23,861 (50.7) | <0.001 | <0.001 |
| CKD | 2183 (25.1) | 27,934 (31.7) | 15,778 (33.5) | <0.001 | <0.001 |
| COPD | 1199 (13.8) | 17,366 (19.7) | 9264 (19.7) | <0.001 | <0.001 |
| Asthma | 1790 (20.6) | 32,231 (36.6) | 18,117 (38.5) | <0.001 | <0.001 |
| Smoking | 1105 (12.7) | 15,167 (17.2) | 7803 (16.6) | <0.001 | <0.001 |
| Liver Disease |  |  |  |  |  |
| Abbreviations: AC, Anticoagulation; CAD, Coronary Artery Disease; CKD, Chronic Kidney Disease; COPD, Chronic Obstructive Pulmonary Disease; DAPT, Dual Antiplatelet Therapy; SAPT, Single Antiplatelet Therapy; SD, Standard Deviation |  |  |  |  |  |
| Comorbidities were identified using ICD-10 codes. Please refer to Supplement for list of ICD-10 codes used to define comorbidities. |  |  |  |  |  |

After 1:1 propensity score matching, 8706 patients in group 1 (AC alone) were matched to 8706 patients in group 2 (AC + SAPT), while 8684 patients in group 1 (AC alone) were matched to 8684 patients in group 3 (AC + DAPT). Comparison on baseline characteristics in propensity matched cohorts are reported in *Table 2,* in which, while some characteristics differed statistically, differences were marginal numerically.

**Table 2:**
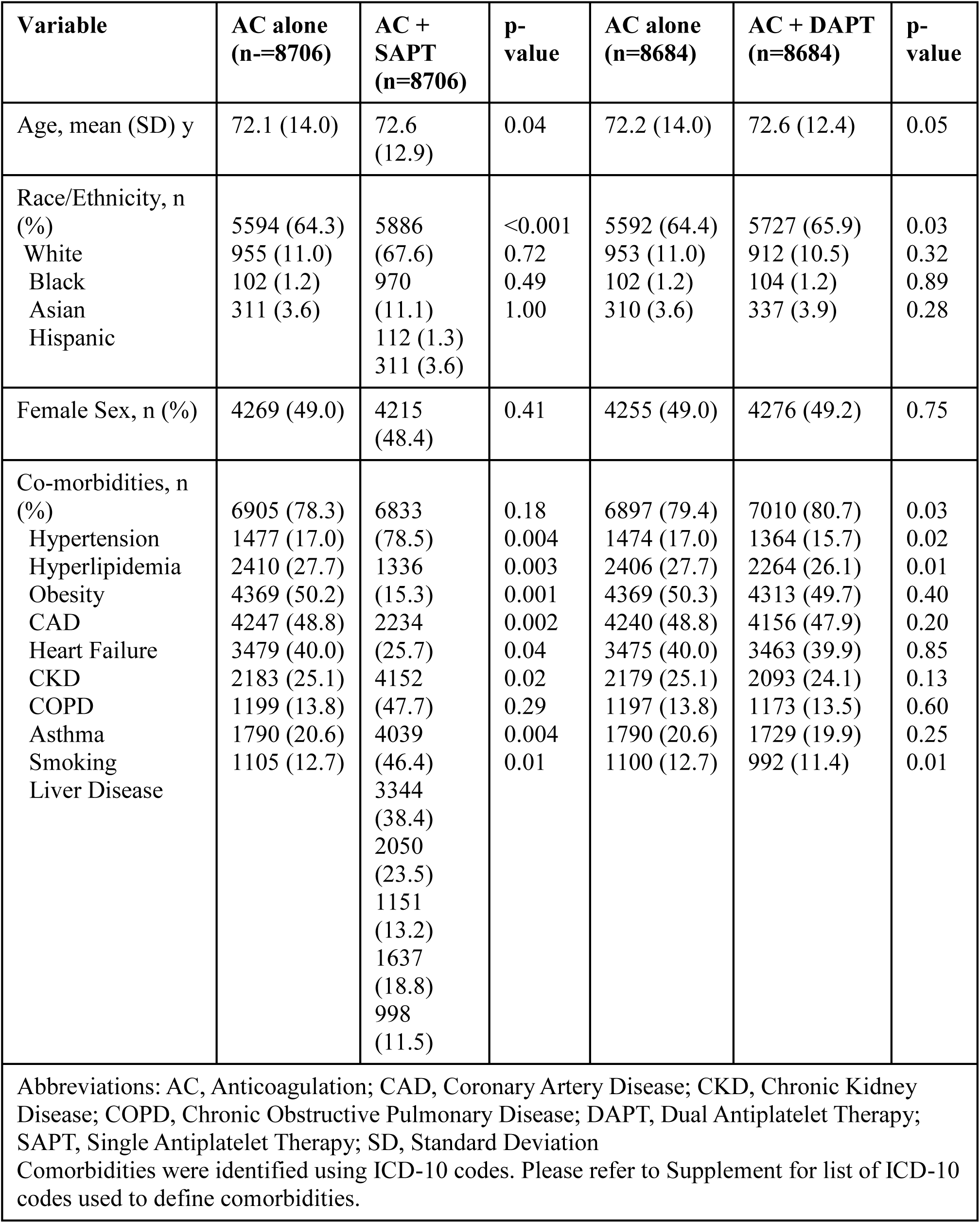
Baseline Characteristics After Propensity-Score Matching.

### Intracerebral Hemorrhage

After 1:1 propensity score matching, 3.7% with AC alone, 4.1% with AC+SAPT and 3.8% with AC+DAPT had a new diagnosis of ICH. By 12 months, 4.4%, 4.9% and 4.9% had a new diagnosis of ICH in groups 1, 2 and 3 respectively. Throughout entire follow-up until query date, 4.9% with AC alone, 6.2% with AC+SAPT and 6.6% with AC + DAPT had a new diagnostic code for ICH (p<0.0001) *(Figure 2)*.

**Figure 2:**
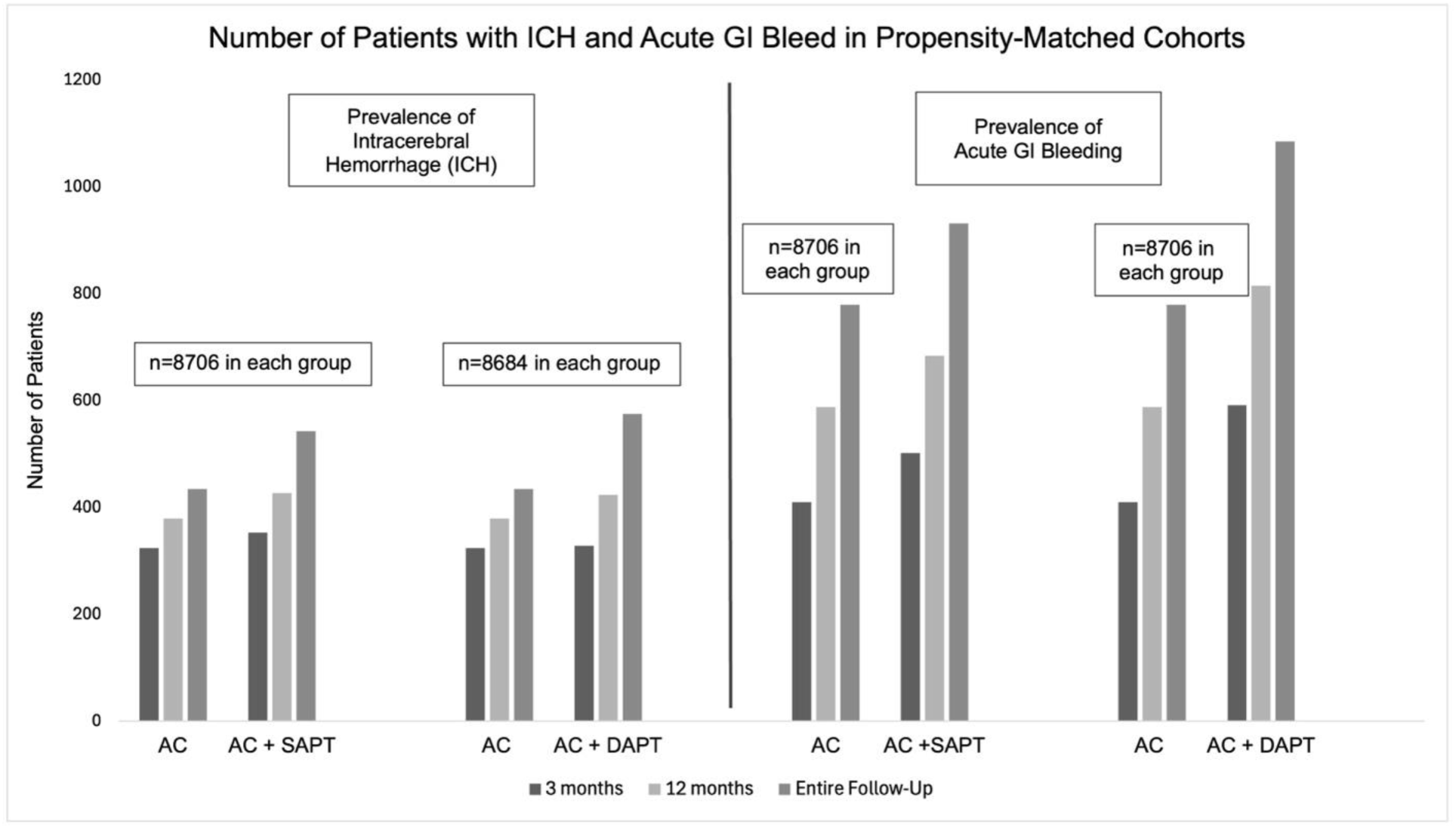
Prevalence of ICH and Acute GI Bleeding Outcomes at Various Time Points after Index Event in Propensity Score Matched Cohorts. This figure shows the number of patients with ICH and acute GI bleeding at 3 months, 12 months and entire follow-up until query date in propensity score matched cohorts of patients with AC, AC + SAPT and AC + DAPT.

In logistic regression after propensity score matching, combining SAPT with AC (Odds Ratio [OR] [95% Confidence Intervals (CI)], 1.09 [0.94-1.28], *p*=0.26) or DAPT with AC (OR [95% CI], 1.01 [0.87-1.18], *p*=0.87) was not associated with increased odds for ICH at 3 months compared to AC alone. Similarly, at 12 months after index event, odds for ICH when combining SAPT with AC (OR [95% CI], 1.13 [0.98-1.31], *p*=0.08) or DAPT with AC (OR [95% CI], 1.13 [0.98-1.30], *p*=0.1) was not increased when compared to AC alone. However, the lifetime odds (through entire follow-up period after index event until query date) for ICH was significantly higher with SAPT + AC (OR [95% CI], 1.26 [1.11-1.44], *p*<0.001) and DAPT + AC (OR [95% CI], 1.34 [1.18-1.53], *p*<0.001), when compared to AC alone. *(Table 3)*.

**Table 3:** Risk of ICH in AC monotherapy vs. combination therapy in propensity score matched cohort.

| **Risk of ICH** | **3-months Odds Ratio (95% CI)** | **P-value** | **12-months Odds Ratio (95% CI)** | **P-value** | **Lifetime Odds Ratio (95%CI)** | **P-value** |
| --- | --- | --- | --- | --- | --- | --- |
| AC+SAPT vs. AC alone (n=8706 per group) | 1.09 (0.94-1.28) | 0.26 | 1.13 (0.98-1.31) | 0.08 | 1.26 (1.11-1.44) | **<0.001** |
| AC+DAPT vs. AC alone (n=8684 per group) | 1.01 (0.87-1.18) | 0.87 | 1.13 (0.98-1.30) | 0.10 | 1.34 (1.18-1.53) | **<0.001** |
| Abbreviations: AC, Anticoagulation; DAPT, Dual Antiplatelet Therapy; SAPT, Single Antiplatelet Therapy; ICH: intracerebral hemorrhage, CI: Confidence Interval |  |  |  |  |  |  |

**Table 4:** Risk of GI bleed in AC monotherapy vs. combination therapy in propensity score matched cohort.

| **Risk of Acute GI Bleeding** | **3-months Odds Ratio (95% CI)** | **p-value** | **12-months Odds Ratio (95% CI)** | **P-value** | **Lifetime Odds Ratio (95% CI)** | **P-value** |
| --- | --- | --- | --- | --- | --- | --- |
| AC+SAPT vs. AC alone<br>(n=8706 per group) | 1.24 (1.08-1.42) | **0.002** | 1.18 (1.05-1.32) | **0.005** | 1.22 (1.10-1.35) | **<0.001** |
| AC+DAPT vs. AC alone<br>(n=8684 per group) | 1.47 (1.29-1.68) | **<0.001** | 1.43 (1.28-1.59) | **<0.001** | 1.45 (1.31-1.60) | **<0.001** |
| Abbreviations: AC, Anticoagulation; DAPT, Dual Antiplatelet Therapy; SAPT, Single Antiplatelet Therapy; GI: gastrointestinal, CI: Confidence Interval |  |  |  |  |  |  |

### Acute GI Bleeding

In contrast to ICH, proportions of patients with acute GI bleeding at all time points were proportionately higher after combination of SAPT or DAPT with AC compared to AC alone at all time points (all p≤0.005) *(Figure 2).* In logistic regression, odds for acute GI bleeding were significantly increased with combination therapy when compared to AC alone at 3 months (SAPT + AC: OR [95% CI], 1.24 [1.08-1.42], *p*=0.002; DAPT + AC: OR [95% CI] 1.47 [1.29-1.68], *p*<0.001). Similarly, at 12 months the odds for acute GI bleeding were significantly higher with SAPT + AC (OR [95% CI], 1.18 [1.05-1.32], *p*=0.005) and DAPT + AC (OR [95% CI], 1.43 [1.28-1.59], *p*<0.001) compared to AC alone. Finally, the lifetime odds of acute GI bleeding with SAPT + AC (OR [95% CI], 1.22 [1.10-1.35], p<0.001) and triple therapy (OR [95% CI], 1.45 [1.31-1.60], *p*<0.001) were also significantly higher compared to AC monotherapy.

## Discussion

In this large multi-year multi-center retrospective cross-sectional observational study using the TriNetX healthcare analytics platform on a cohort of >140,000 AIS patients with concomitant diagnoses of acute MI and AF across multiple US centers, 94% received combination therapy with antiplatelets and AC, and >30% were prescribed triple antithrombotic therapy. Combining SAPT or DAPT with AC was not associated with increased risk of non-traumatic ICH in the first 3-12 months but was associated with a significantly higher ICH risk in the long-term follow-up > 12 months within TriNetX. On the contrary, risk of acute GI bleeding was significantly higher at 3 months, 12 months, and throughout follow-up. These findings suggest that combining SAPT or DAPT with AC may not be associated with increased risk of ICH in the first 3-12 months, but this risk may increase significantly in the long-term indicating that such therapy should be limited to shorter durations in AIS patients. On the contrary, our study also demonstrated that a higher risk of GI bleeding exists as early as the first 3 months after initiation and continues in the long-term, with greater risk with triple therapy, suggesting that early initiation of preventative strategies to mitigate GI bleeding, such as H2-receptor blockers or proton pump inhibitors, may be warranted when combining antiplatelets with AC in AIS patients.

### Strengths and Limitations

The strengths of this study include data from the largest cohort to date of AIS patients with comorbid AF and acute MI from more than 50 US academic and affiliated community health centers included in the TriNetX database. TriNetX, a federal healthcare analytics platform, has been validated for comparative effectiveness research and provides the benefit of large volumes of patient data, including diagnostic and medication codes, by collating data from electronic health records across large number of US centers in real-time. Moreover, our study also provides the largest propensity-score matched analysis of AIS patients in the contemporary literature that were prescribed antiplatelets in combination with AC, particularly triple antithrombotic therapy, providing important insights into prescription practices for this important clinical conundrum. Finally, our study analyzed bleeding outcomes at multiple longitudinal time points, elucidating potential safety or harm of combination antiplatelet and AC therapy after AIS in the short- and long-term.

Important limitations of this study need to be highlighted. First, adjudication of cases and outcomes were done using ICD-10-CM codes, which may be prone to misclassification biases as they rely on coders and clinicians to assign appropriate diagnostic codes. For example, MI in the past may be coded as acute MI upon admission. However, separate ICD-10-CM codes exist for acute versus chronic MI, acute versus chronic strokes, traumatic versus non-traumatic ICH, and only specific ICD-10-CM codes for acute disease processes (Methods and Supplement) were used in this study. Moreover, our very large sample size may, at least in part, mitigate the risk of this bias. Second, while we used propensity-score matching to balance cohorts for multiple vascular risk factors and demographic characteristics that are known to influence bleeding outcomes, we were unable to account for important known confounders due to lack of availability of these data in TriNetX, such as admission stroke severity, imaging characteristics such as stroke size, cerebral microbleeds, small vessel disease, intra- or extra-cranial atherosclerosis, left atrial or ventricular thrombi, all of which may influence the use of combination antithrombotic therapy, their duration, and the risk of long-term bleeding. Similarly, the retrospective nature of this study also limits ability to adjust for all possible unknown confounders, limiting causal interpretation. Third, an important limitation of our analysis, is the lack of details regarding the duration and dosing of medication prescription. While all patients received AC plus antiplatelets when entering the dataset, data on discontinuation of AC or antiplatelet agents in the future was not available, thus we were unable to delineate how long these combination therapies were prescribed, limiting determination of a temporal relationship and hence, assigning a causal interpretation to our findings. We therefore intentionally did not compare the bleeding risk between AC plus SAPT versus AC plus DAPT groups as many patients could have transitioned from DAPT to SAPT at some point in the course of therapy making the assessment between the 2 groups complex and unreliable. Forth, we did not examine the association with short- and long-term ischemic outcomes due to limited ability to identify new diagnoses of recurrent stroke and the lack of neuroimaging data within TriNetX. Nonetheless, the large number of patients in our cohorts, propensity-score matching, and availability of long-term follow-up data provide important insights in this specific patient population, given limited availability of data in current literature. Validation of our findings in prospective studies and randomized clinical trials are necessary. Finally, TriNetX does not allow access to individual patient data, but its built-in statistical method for propensity score matching uses the greedy nearest neighbor algorithm. Other approaches, such as overlap propensity score or inverse probability of treatment weighting, may yield different outcomes; however, we were unable to evaluate these methods.

### Implications of the Study

Several clinical trials have examined the risk of bleeding when combining AC with SAPT or DAPT in the cardiac patient population. The PIONEER AF-PCI randomized 2124 patients with non-valvular AF who needed PCI with stent placement for acute coronary syndrome (ACS) and or stable ischemic heart disease, to receive rivaroxaban in combination with clopidogrel, very low dose rivaroxaban with clopidogrel and aspirin or standard therapy that included warfarin (INR 2-3), clopidogrel, and aspirin for 12 months (Gibson, 2016). The primary outcome of clinically significant bleeding over the treatment period of 12 months was lower in the groups receiving rivaroxaban and clopidogrel compared to triple therapy with warfarin, clopidogrel, and aspirin. The RE-DUAL PCI, a multicenter noninferiority trial, randomized 2725 patients with AF and recent PCI for ACS or stable ischemic heart disease, to receive dabigatran plus clopidogrel versus triple therapy with warfarin, clopidogrel and aspirin (Cannon 2017). Major bleeding or clinically relevant nonmajor bleeding, was overall lower in AC and SAPT (15.4%) compared to triple therapy (26.9%) (HR, 0.72; 95% CI, 0.58 to 0.88; P<0.001). Similarly, the ENTRUST-AF PCI non-inferiority trial randomized 1506 patients with AF receiving PCI for ACS or stable ischemic heart disease to receive edoxaban plus clopidogrel versus triple therapy with warfarin, clopidogrel and aspirin (Vranckx 2019). Edoxaban plus clopidogrel was non-inferior to triple therapy with respect to clinically significant bleeding (17% versus 20%) without an increase in ischemic events. Finally, the AUGUSTUS trial randomized 4614 patients with AF and recent ACS or PCI to 4 groups: apixaban plus clopidogrel, apixaban plus aspirin and clopidogrel, warfarin and clopidogrel or warfarin, aspirin and clopidogrel (Lopes 2019). Primary outcome was defined as major or clinically relevant nonmajor bleeding. Apixaban when compared to warfarin was associated with lower rates of major or clinically relevant non-major bleeding regardless of prior history of stroke. This study concluded that anticoagulation with a DOAC and a P2Y12 is likely appropriate in the majority of patients and triple therapy with aspirin may be contributing to bleeding risks. More importantly, AUGUSTUS highlights that avoiding aspirin resulted in a 47% lower risk of bleeding than adding aspirin without a significantly higher incidence of coronary ischemic events. Collectively, these trials demonstrate that SAPT and DOAC may be safer with respect to bleeding outcomes when compared to triple antithrombotic therapy in patients with AF and CAD. Unfortunately, with the exception of the AUGUSTUS trial, all the remaining trials intentionally excluded patients with a recent diagnosis of stroke. Moreover, in the AUGUSTUS trial only 14% of the participants had a history of stroke or TIA, making it challenging to extrapolate these study findings for the purpose of secondary stroke prevention in patients presenting with a diagnosis of AIS. Our study thus provides novel insight into the practices around combining antiplatelets with AC in AIS patients with AF and ACS in current clinical practice and delineates the short and long-term risk of bleeding in this specific patient population.

In a recent study of 93 patients receiving triple antithrombotic therapy, readmission rate due to bleeding was 2.2% at 14 days and 6.5% at 30 days (Robichaux 2024). Importantly, the indication for antiplatelet therapy in this study was not limited to acute coronary syndrome, and ∼15% received it for a diagnosis of AIS. Additionally, indication for AC was AF in 40% of the patients but, VTE and other indications were also included. Within 30 days of initiation of triple therapy, 4.3% of patients endured major bleeding as defined by the International Society on Thrombosis and Hemostasis. These proportions appear to be similar to our study, which also showed an average risk of ICH and acute GI bleeding of 4% and 6% respectively in patients receiving SAPT or DAPT and AC after AIS.

## Conclusions and Future Research

This large multicenter retrospective cross-sectional observational cohort study demonstrates that antiplatelets are commonly combined with anticoagulation in AIS patients with concomitant AF and acute MI. Within the limitations of this retrospective electronic health records-based registry study, we did not identify an increased risk of ICH with addition of antiplatelets to AC in AIS patients with concomitant AF and acute MI in the short term (3 months); however, we observed a 25% increase in long-term odds of ICH after adding 1 antiplatelet agent and a 35% increase after adding 2 antiplatelet agents. On the other hand, risk of acute GI bleeding was increased in the short and long-term after adding antiplatelet therapy to AC therapy, with a 20% increase in odds with 1 antiplatelet and 45% increase in odds with 2 antiplatelet agents. This highlights the fact that the additional risk of triple therapy or SAPT and AC over AC monotherapy should be taken into consideration when making decisions about adding antiplatelet medications in AIS patient with AF and concomitant acute MI. Larger prospective cohort studies and randomized clinical trials are warranted to validate our findings and study the safety and efficacy of combining antiplatelet agents with AC for secondary stroke prevention in AIS survivors with history of AF and concomitant ACS/CAD.

## Data Availability

The data is available and accessible on TrinetX.

## Non-standard Abbreviations and Acronyms

AF: Atrial fibrillation
AC: Anticoagulation
ASCVD: Atherosclerotic cardiovascular disease
DAPT: Dual anti-platelet therapy
ICH: Intra-cerebral hemorrhage
PCI: Percutaneous coronary intervention
SAPT: Single anti-platelet therapy
SIHD: Stable ischemic heart disease

## Acknowledgement

The authors are grateful to William Merwin M.D. who contributed to editing and formatting of this publication.

---

**Supplementary Table 1:** ICD-10 Codes and associated disease states.

| Disease State | ICD-10 CM Code |
| --- | --- |
| Hypertension | I10 |
| Type 2 Diabetes | E11 |
| Heart Failure | I50 |
| Coronary Artery Disease | I25.10 |
| Chronic Obstructive Pulmonary Disease | J44.9 |
| Asthma | J45.909 |
| Smoking (Tobacco use disorder) | F17.210 (current) |
| Chronic Kidney Disease | N18.9 |
| Obesity | E66.9 |
| Hyperlipidemia | E78.5 |

